# Peginterferon Lambda-1a for treatment of outpatients with uncomplicated COVID-19: a randomized placebo-controlled trial

**DOI:** 10.1101/2020.11.18.20234161

**Authors:** Prasanna Jagannathan, Jason R. Andrews, Hector Bonilla, Haley Hedlin, Karen B. Jacobson, Vidhya Balasubramanian, Natasha Purington, Savita Kamble, Christiaan R. de Vries, Orlando Quintero, Kent Feng, Catherine Ley, Dean Winslow, Jennifer Newberry, Karlie Edwards, Colin Hislop, Ingrid Choong, Yvonne Maldonado, Jeffrey Glenn, Ami Bhatt, Catherine Blish, Taia Wang, Chaitan Khosla, Benjamin A. Pinsky, Manisha Desai, Julie Parsonnet, Upinder Singh

## Abstract

Type III interferons have been touted as promising therapeutics in outpatients with coronavirus disease 2019 (COVID-19). We conducted a randomized placebo-controlled trial in 120 patients with mild to moderate COVID-19 to determine whether a single, 180 mcg subcutaneous dose of Peginterferon Lambda-1a (Lambda) could shorten the duration of viral shedding (primary endpoint) or symptoms (secondary endpoint, NCT04331899). In both the 60 patients receiving Lambda and the 60 receiving placebo, the median time to cessation of viral shedding was 7 days (hazard ratio [HR] = 0.81; 95% confidence interval [CI] 0.56 to 1.19). Symptoms resolved in 8 and 9 days in Lambda and placebo, respectively (HR 0.94; 95% CI 0.64 to 1.39). At enrollment; 41% of subjects were SARS-CoV-2 IgG seropositive; compared to placebo, lambda tended to delay shedding cessation in seronegatives (aHR 0.66, 95% CI 0.39-1.10) and to hasten shedding cessation in seropositives (aHR 1.58, 95% CI 0.88-2.86; p for interaction = 0.03). Liver transaminase elevations were more common in the Lambda vs. placebo arm (15/60 vs 5/60; p = 0.027). In this study, a single dose of subcutaneous Peginterferon Lambda-1a neither shortened the duration of SARS-CoV-2 viral shedding nor improved symptoms in outpatients with uncomplicated COVID-19.

---

Coronavirus Disease 2019 (COVID-19), caused by the Severe Acute Respiratory Syndrome Coronavirus 2 (SARS-CoV-2), has led to nearly 1 million deaths worldwide as of September 2020.^1^ Although most infected patients display mild symptoms, even uncomplicated infections can contribute to transmission to those with co-morbid conditions and other high risk groups, increasing overall mortality.^2^ With the unprecedented health and economic threats imposed by COVID-19, therapeutics are urgently needed to shorten the duration of viral shedding, relieve symptoms, and prevent hospitalizations..

Interferons (IFNs) are promising anti-SARS-CoV-2 therapeutics, given their importance in the early response to viral infections.^3^ Innate immune sensing of viral nucleic acids leads to production of type I (IFN-α, IFN-β) and type III (IFN-λ) IFNs that, after binding to their cognate receptors, activate genes critical for host protection.^4-6^ SARS-CoV-2 encodes proteins that suppress production of endogenous IFN^7^, and infection has been associated with markedly reduced type I and type III IFN signaling^8^, particularly in patients with severe manifestations of disease.^9,10^ Both type I and type III IFNs inhibit SARS-CoV-2 *in vitro*^11,12^, suggesting potential utility of exogenous IFN administration to aid in viral control and prevent disease progression. In support of this hypothesis, recent trials in hospitalized COVID-19 patients have reported that type I IFN administration may reduce the duration of viral shedding and symptoms.^13-15^

Whereas cognate receptors to type I IFNs are expressed ubiquitously, the receptor complex (IL28R) for IFN-λ is expressed on only a few cell types, including epithelial cells in the gastrointestinal and respiratory tracts.^4,16,17^ These cellular affinities have led investigators to use this agent to target viral hepatitides^18,19^ and respiratory viral infections^16^. In a murine model of influenza infection, IFN-λ treatment post-infection was associated with significantly lower mortality compared to mice treated with IFN-α, and this was associated with lower influenza viral loads^20^. A pegylated form of recombinant IFN-λ, Peginterferon Lambda-1a (Lambda) has been developed for the treatment of chronic viral hepatitis. Lambda, given weekly as 180 mcg subcutaneous injections, has comparable antiviral efficacy and an improved tolerability profile compared with type I IFN for the treatment of hepatitis^21^, likely due to its relatively limited receptor distribution. In a murine model of SARS-CoV-2 infection, subcutaneous administration of Lambda prophylactically or early after infection diminished SARS-CoV-2 replication in the lower respiratory tracts of mice *in vivo*^11^.

Lambda has thus emerged as a promising treatment candidate for SARS-CoV-2^22,23^ given a plausible mechanism of action, the suppression of IFN activity by SARS-CoV2, and *in vitro* and *in vivo* studies showing that IFN-λ administration can inhibit SARS-CoV-2 replication. To date, no therapies have been approved for outpatients with mild to moderate COVID-19 disease. We therefore conducted a randomized, placebo-controlled trial of Lambda for outpatients with uncomplicated SARS-CoV-2 infection. We tested the hypothesis that a single, 180 mcg subcutaneous injection of Lambda would be associated with a shorter duration of viral shedding in comparison to a normal saline placebo injection.

## Results

### Cohort Characteristics

We enrolled 120 participants between April 25 and July 17, 2020, of whom 110 (91.7%) completed 28 days of follow up (**Fig 1a**). The median age was 36 years (range 18-71), 50 participants (41.7%) were female, and 75 (62.5%) were Latinx ethnicity (**Table 1**). Eight (6.7%) participants were asymptomatic at baseline. Of those with symptoms, the median duration of symptoms prior to randomization was five days. The most common symptoms were fatigue, cough, headache, and myalgias (**Table 1, Fig S1**). Only 13 (10.8%) participants had an elevated temperature (>99.5°F) at baseline; the median oxygen saturation was 98%.

**Table 1.** Baseline characteristics of study participants.

|  | Treatment arm |  | Overall<br>(N=120) | ASD |
| --- | --- | --- | --- | --- |
|  | Lambda<br>(N=60) | Placebo<br>(N=60) |  |  |
| **Age in years, median (range)** | 37 (18-66) | 34 (20-71) | 36 (18-71) | 0.14 |
| **Female, n (%)** | 24 (40.0%) | 26 (43.3%) | 50 (41.7%) | 0.07 |
| **Race / Ethnicity, n (%)** |  |  |  | 0.46 |
| Latinx | 34 (56.7%) | 41 (68.3%) | 75 (62.5%) |  |
| White | 18 (30.0%) | 15 (25.0%) | 33 (27.5%) |  |
| Asian | 3 (5.0%) | 4 (6.7%) | 7 (5.8%) |  |
| Native Hawaiian or other Pacific Islander | 2 (3.3%) | 0 (0%) | 2 (1.7%) |  |
| Unknown | 2 (3.3%) | 0 (0%) | 2 (1.7%) |  |
| More than one race | 1 (1.7%) | 0 (0%) | 1 (0.8%) |  |
| **BMI (kg/m<sup>2</sup>), median (IQR)** | 27.6 (25.4-31.1) | 28.5 (24.8-32.3) | 27.7 (24.9-32.0) | 0.06 |
| **Asymptomatic at baseline, n (%)** | 6 (10.0%) | 2 (3.3%) | 8 (6.7%) | 0.27 |
| **Duration of symptoms in days prior to randomization, median (IQR)<sup>1</sup>** | 4 (3-6) | 5 (3-5) | 5 (3-6) | 0.13 |
| **Symptoms at baseline, n (%)** |  |  |  |  |
| Fatigue | 33 □ (55%) | 42 □ (70%) | 75 □ (62.5%) | 0.25 |
| Cough | 33 (55.0%) | 36 (60.0%) | 69 (57.5%) | 0.08 |
| Headache | 29 □ (48.3%) | 36 □ (60%) | 65 □ (54.2%) | 0.22 |
| Myalgias | 29 □ (48.3%) | 34 □ (56.7%) | 63 □ (52.5%) | 0.15 |
| Decreased taste or smell | 25 □ (41.7%) | 32 □ (53.3%) | 57 □ (47.5%) | 0.22 |
| Chills | 22 □ (36.7%) | 27 □ (45%) | 49 □ (40.8%) | 0.16 |
| Sore throat | 22 □ (36.7%) | 23 □ (38.3%) | 45 □ (37.5%) | 0.04 |
| Joint pain | 19 □ (31.7%) | 19 □ (31.7%) | 38 □ (31.7%) | <0.001 |
| Diarrhea | 16 □ (26.7%) | 18 □ (30%) | 34 □ (28.3%) | 0.06 |
| Nausea | 11 □ (18.3%) | 23 □ (38.3%) | 34 □ (28.3%) | 0.44 |
| Shortness of breath | 16 (26.7%) | 16 (26.7%) | 32 (26.7%) | 0.01 |
| Chest pain/pressure | 14 □ (23.3%) | 13 □ (21.7%) | 27 □ (22.5%) | 0.05 |
| Runny nose | 10 □ (16.7%) | 16 □ (26.7%) | 26 □ (21.7%) | 0.23 |
| Abdominal pain | 7 □ (11.7%) | 7 □ (11.7%) | 14 □ (11.7%) | <0.001 |
| Rash | 4 □ (6.7%) | 5 □ (8.3%) | 9 □ (7.5%) | 0.06 |
|  | Lambda<br>(N=60) | Placebo<br>(N=60) |  |  |
| Vomiting | 1 □ (1.7%) | 5 □ (8.3%) | 6 □ (5%) | 0.31 |
| **Vital signs at enrollment** |  |  |  |  |
| Temperature 99.5F+, n (%) | 6 (10.0%) | 7 (11.7%) | 13 (10.8%) | 0.05 |
| Oxygen saturation, median (IQR) | 98 (2.5) | 99 (3) | 98 (3) |  |
| **Baseline laboratory values, median (IQR)** |  |  |  |  |
| White blood cell (WBC) count, cells/μl | 5.5 (4.3-6.8) | 5.6 (4.0-7.5) | 5.5 (4.1-7.1) | 0.08 |
| Absolute lymphocyte count (ALC), cells/μl | 1.5 (1.2-1.9) | 1.5 (1.2-2.3) | 1.5 (1.2-2.2) | 0.14 |
| Aspartate aminotransferase, IU/L | 31 (26-41) | 30 (25-39.3) | 30 (25-41) | 0.25 |
| Alanine aminotransferase, IU/L | 32.5 (21-52.3) | 30.5 (23-47.5) | 31.5 (22-50.3) | 0.25 |
| **Baseline oropharyngeal SARS-CoV-2 cycle threshold, median (IQR) <sup>2</sup>** | 30.9 (26.4-33.8) | 29.3 (26.4-34.3) | 30.3 (26.4-34.3) | 0.12 |
| **Baseline Log10 Viral Load, median (IQR) <sup>2</sup>** | 4.2 (3.3 - 5.5) | 4.7 (3.2 - 5.5) | 4.4 (3.2 - 5.5) | 0.12 |
| **Baseline SARS-CoV-2 IgG seropositivity, n (%)** | 21 (35.0%) | 28 (46.7%) | 49 (40.8%) | 0.24 |
| **Sum of risk factors, median (IQR)** | 3 (2-3) | 3 (2-4) | 3 (2-3) | 0.07 |

IQR = inner quartile range; ASD = absolute standardized difference. sum of risk factors is defined as the number of relevant severe disease risk factors present at baseline (presence of either temperature of 99.5F+, cough, or shortness of breath; age 60+; male sex; Black race; Latinx ethnicity; BMI 30+; ALC<1000; ALT 94+).
among n=103 participant who reported symptoms prior to randomization (n=48 in lambda and n=55 in placebo).
among n=87 participants with detectable OP virus (n=44 in lambda and n=43 in placebo).

**Figure 1.**
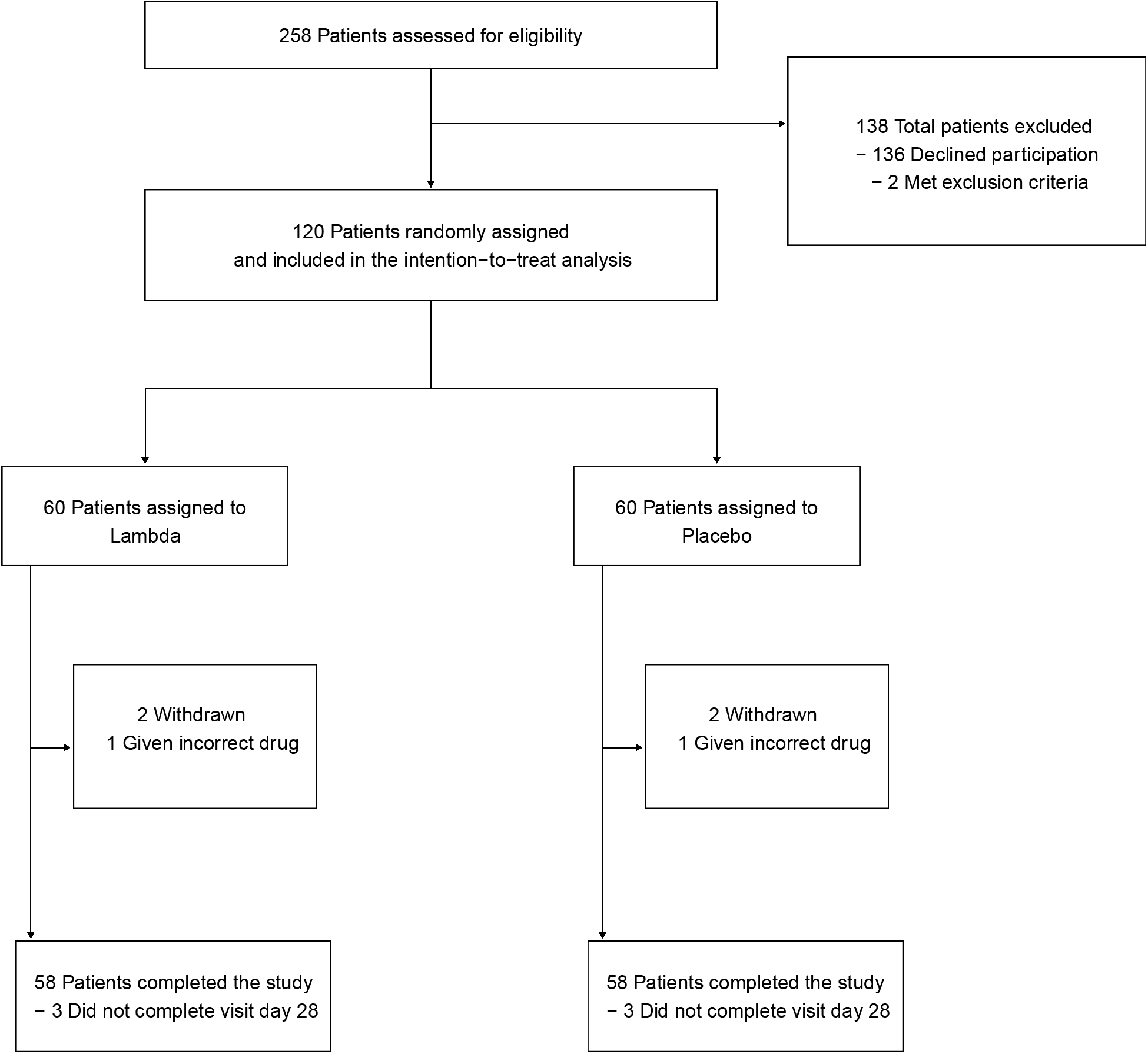
CONSORT Diagram

### Baseline oropharyngeal viral RNA level and SARS-CoV-2 IgG serostatus

The presence of SARS-CoV-2 RNA was assessed at baseline and at 8 follow-up visits using oropharyngeal swabs, a Centers for Disease Control approved method for SARS-CoV-2 detection^24^. This method was selected given the frequency of repeated assessments and improved tolerability for participants compared with nasopharyngeal swabs. The median SARS-CoV-2 oropharyngeal viral PCR cycle threshold at enrollment was 30.3 (corresponding to a median viral load of 4.4 Log10 copies/ml), and did not differ significantly between groups (**Table 1**).

IgG antibody titers against the SARS-CoV-2 spike receptor binding domain (RBD) were assessed at enrollment by enzyme linked immunosorbent assay.^25^ At enrollment, 49 (40.8%) participants were SARS-CoV-2 IgG seropositive. Baseline SARS-CoV-2 serostatus did not significantly differ between groups (**Table 1**). The median duration of symptoms prior to enrollment was significantly shorter in SARS-CoV-2 IgG seronegative vs. seropositive participants (median [interquartile range [IQR]]: 3.5 [2.5-5] days vs. 5 [4-7] days, P=0.0051). Seronegative participants also had significantly higher oropharyngeal viral RNA levels at enrollment compared with seropositive participants (median [IQR] log_10_ viral load 4.4 [2.5] vs. 2.0 [2.4]).

### Primary Virologic Analysis

Of 120 enrolled participants, 60 were randomized to receive Lambda and 60 randomized to receive placebo and included in the analysis. The median time to cessation of oropharyngeal viral shedding was 7 days in both arms (95% CI 5-10 days for placebo vs 5-13 days for Lambda, **Table 2**). There was no significant difference in the adjusted hazard ratio (aHR) for shedding cessation between Lambda and placebo; participants in the lambda arm were 19% less likely to cease shedding at any point during the study period compared to participants in the placebo arm (aHR 0.81, 95% confidence interval [CI] 0.56 to 1.19; p = 0.29, **Fig 2A**.) Overall, 108 participants met the primary endpoint and were not censored. Because two participants, after randomization, inadvertently were injected with the incorrect syringe, we also conducted an as-treated analysis according to treatment actually received. Findings from an as-treated analysis (aHR 0.83, 95% CI 0.56 to 1.21; p = 0.33) and an analysis performed in symptomatic patients only (aHR 0.77, 95% CI 0.52 to 1.15, p = 0.21) were similar.

**Table 2.**
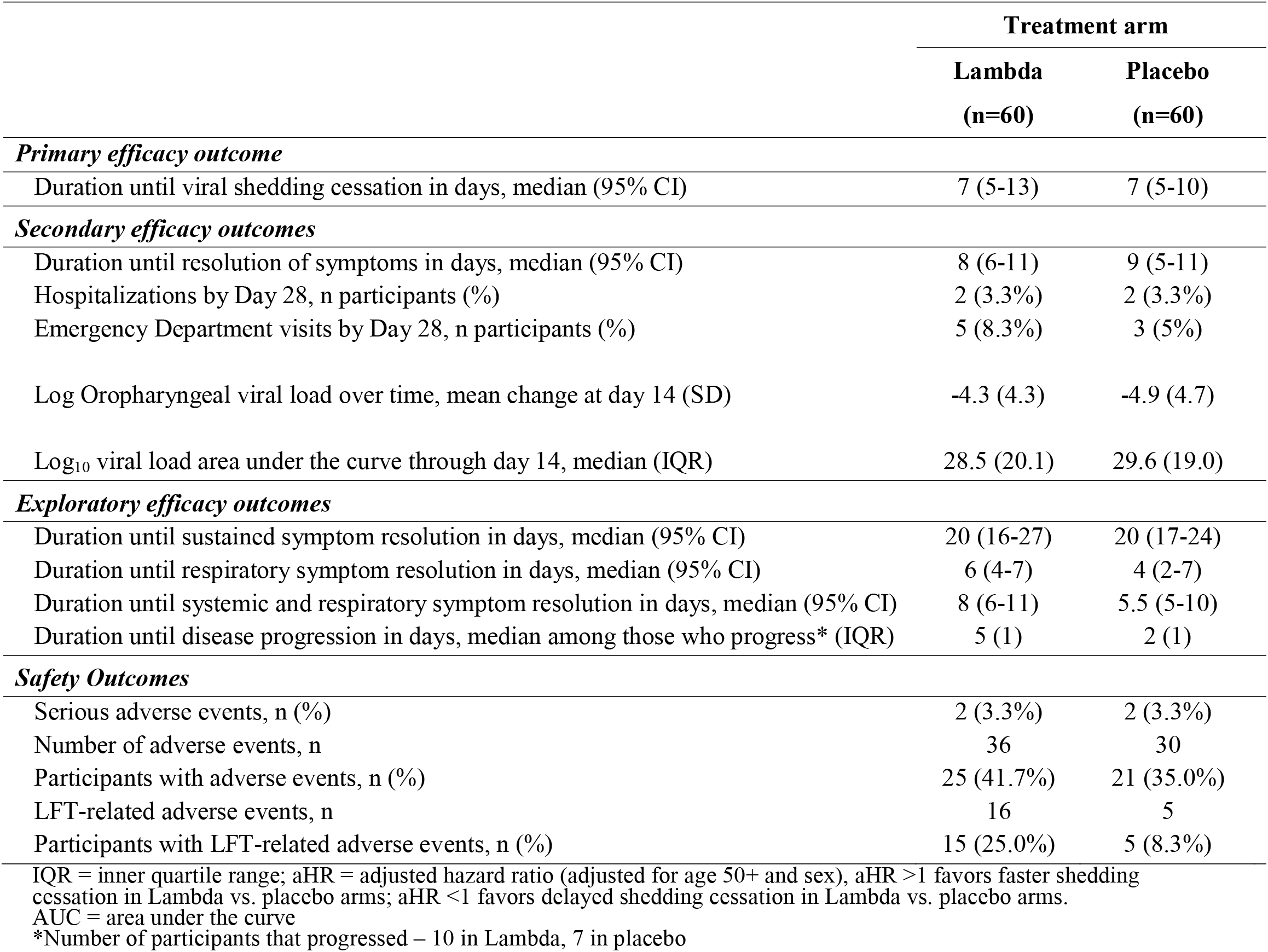
Efficacy and Safety Outcomes.

**Figure 2.**
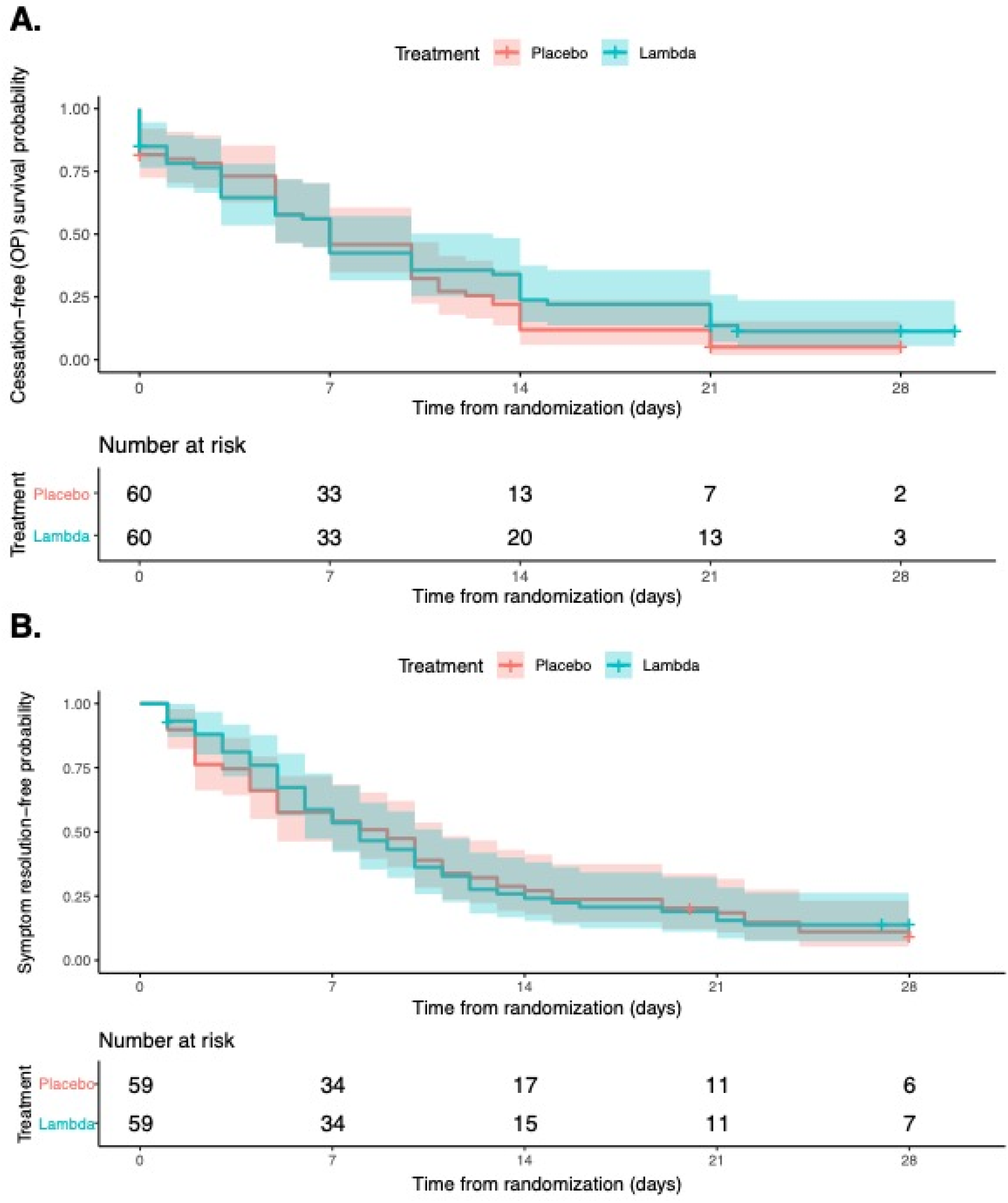
Kaplan-Meier Analyses of the Primary and Key Secondary Outcome in the Intention-to-Treat Population. (A) Time until cessation of SARS-CoV-2 viral shedding from oropharyngeal swabs stratified by treatment arm, Lambda (blue) vs. placebo (red). (B) Time until resolution of all symptoms stratified by treatment arm, Lambda (blue) vs. placebo (red)

In exploratory analysis, seropositivity at baseline was associated with significantly hastened shedding cessation. The median time to shedding cessation was 10 days in SARS-CoV-2 IgG seronegative individuals vs. 3 days in seropositive individuals (95% CI 7-14 days for seronegative vs 1-6 days for seropositive, aHR 2.65, 95% CI 1.74-4.03, P<0.001, **Fig S2**). Baseline serostatus also significantly modified the effect of treatment on time to shedding cessation (p = 0.03). Among seronegative individuals, Lambda delayed shedding cessation compared with placebo (aHR 0.66, 95% CI 0.39-1.10, **Fig 3a**). Among seropositive individuals, Lambda hastened shedding cessation (aHR 1.58, 95% CI 0.88-2.86, **Fig 3a**).

**Figure 3.**
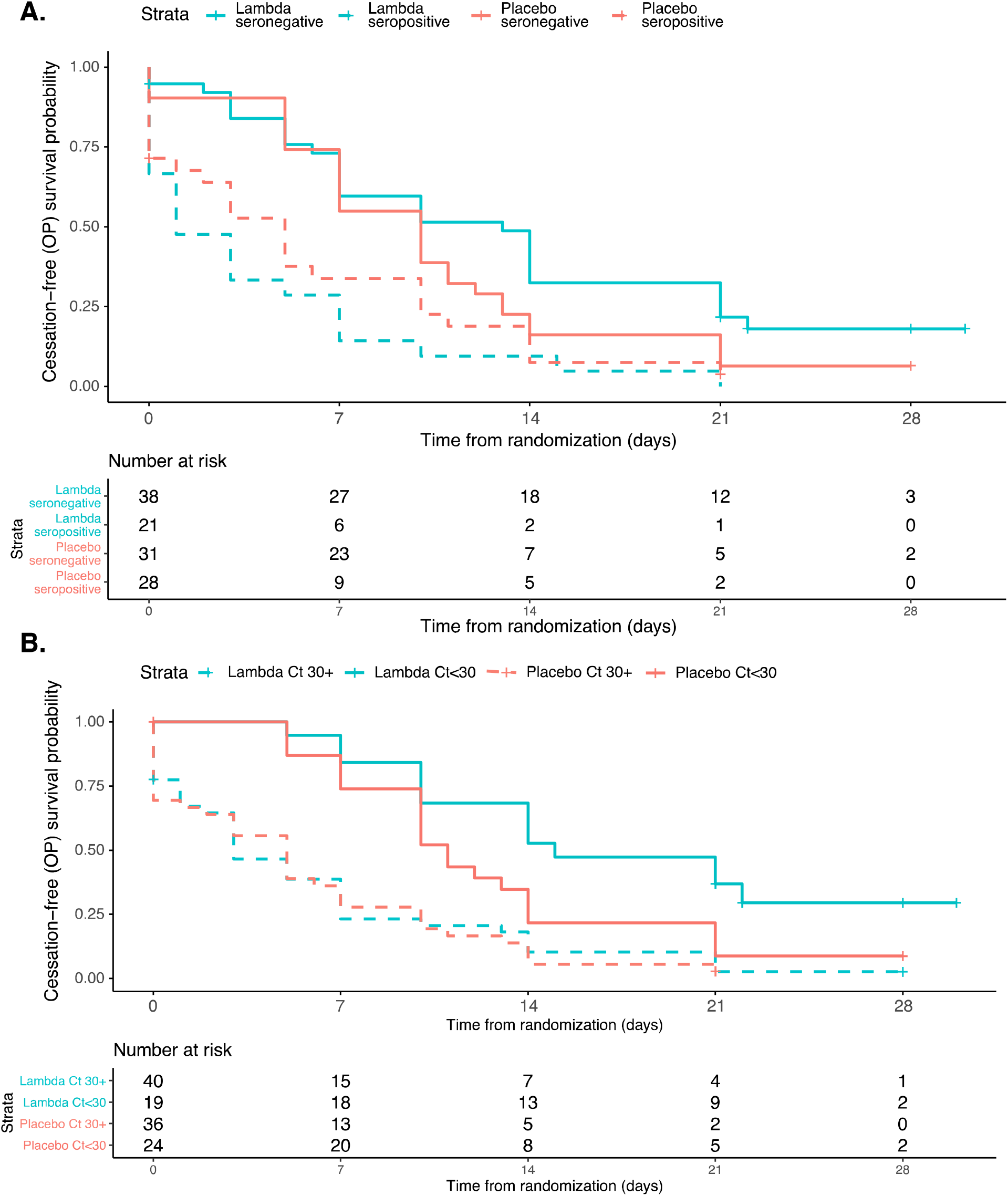
Kaplan-Meier Analyses of the Primary Outcome, Stratified by Baseline Seropositivity and Viral Load. (A) Time until cessation of SARS-CoV-2 viral shedding from oropharyngeal swabs stratified by baseline SARS-CoV-2 seropositivity, seropositive (dashed) and seronegative (solid), and treatment arm, Lambda (blue) vs. placebo (red). (B) Time until cessation of SARS-CoV-2 viral shedding from oropharyngeal swabs stratified by baseline SARS-CoV-2 oropharyngeal virus CT value, CT value >=30 (dashed) and CT value <30 (solid), and treatment arm, Lambda (blue) vs. placebo (red).

Higher oropharyngeal viral RNA levels at baseline were associated with significantly delayed shedding cessation (aHR 0.32 comparing baseline CT <30 vs baseline CT >=30, 95% CI 0.21-0.50, P<0.001, **Fig S3**). Although baseline oropharyngeal viral RNA levels did not significantly modify the effect of treatment on time to shedding cessation (p=0.15), among individuals with high baseline viral RNA levels, Lambda delayed shedding cessation compared with placebo (aHR 0.51, 95% CI 0.26-1.04, **Fig 3b**). There was no difference in shedding cessation between arms with low baseline viral RNA levels (aHR 0.95, 95% CI 0.60-1.52, **Fig 3b**). No other baseline features of interest significantly modified the effect of treatment and the primary outcome (**Table S1)**.

### Secondary analyses

No significant difference in time to resolution of symptoms (aHR 0.94; 95% CI 0.64 to 1.39; p = 0.76, **Fig 2B**) or sustained resolution of symptoms (aHR 0.92; 95% CI 0.60 to 1.41; p = 0.70) was observed, nor did we find any significant difference in resolution of symptom complexes (**Table 2, Fig S4**). Time to clinical progression was not significantly different between the two arms (aHR 1.38; 95% CI 0.52 to 3.63; p = 0.52). Trajectory of viral RNA levels did not vary by treatment arm (p=0.91, **Fig S5**) nor did viral RNA area under the curve (p = 0.95, **Table 2**).

### Adverse Events

Twenty-five (42%) Lambda and 21 (35%) placebo participants experienced adverse events (Table 3). Two serious adverse events (hospitalizations) were reported in each arm. Liver transaminase elevations were more common in the Lambda vs. placebo arm (15 vs 5; p = 0.027). Furthermore, we observed significant elevations in alanine transaminase levels from day 0 to day 5 among individuals randomized to Lambda, but not among individuals randomized to placebo (**Fig 4**). However, there were no associated symptoms and lab abnormalities were not sustained.

**Figure 4.**
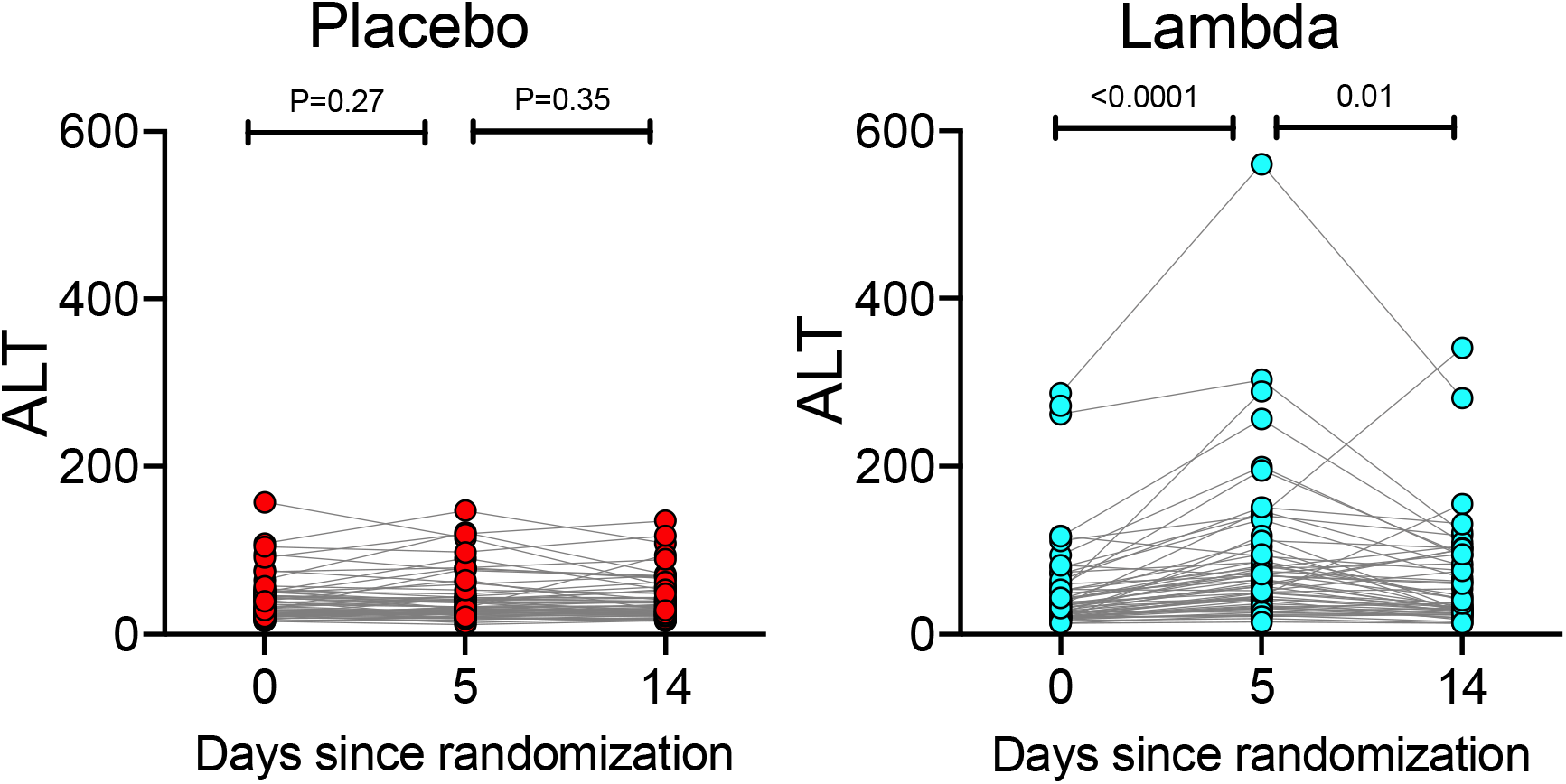
Alanine transaminase levels measured at day 0 and day 5 and 14 post-randomization in Placebo (red) and Lambda (blue) arms.

## Discussion

In this study of outpatients with uncomplicated SARS-CoV-2 infection, a single subcutaneous injection of Lambda did not significantly reduce time to viral clearance or resolution of symptoms compared with placebo. We recruited participants within 72 hours of diagnosis, giving us an excellent opportunity to intervene early within the course of infection. We attained excellent follow-up and retention, with few missed visits (<5%), and little missing data. Despite these strengths and compelling preclinical data--i.e., a plausible mechanism of action, the suppression of IFN activity by respiratory coronaviruses, and both *in vitro* and *in vivo* studies showing inhibition of SARS-CoV-2 replication by IFN-λ^11,12^--this phase 2 trial yielded little promise of efficacy at the tested dose and administration schedule. Lambda was well-tolerated, with few adverse effects, though asymptomatic liver transaminase elevations occurred more frequently in participants randomized to Lambda and are consistent with previous reports.^21,26^

The lack of effect of Lambda was surprising given recently described *in vitro* data and benefits seen in an *in vivo* model with early therapeutic and prophylactic administration^12^. There are several potential reasons for this lack of benefit. First, although we attempted to randomize participants as soon as possible after the COVID-19 diagnosis was made, the median symptom duration was 5 days at the time of randomization, and 40% of participants were already SARS-CoV-2 IgG positive at enrollment. It is possible that earlier administration, or prophylactic administration prior to established infection, would have been beneficial. Arguing against this, we observed no evidence of benefit among SARS-CoV-2 seronegative individuals, who presumably have been infected a shorter period of time. Second, a single, 180 mcg subcutaneous injection of Lambda may not achieve adequate therapeutic levels of drug in the upper respiratory epithelia. Consistent with this, a murine model of SARS-CoV-2 infection found that subcutaneous administration of Lambda did not result in significant reductions of SARS-CoV-2 viral titers in upper respiratory epithelium.^12^ It is possible that higher, or more frequent, dosing may have been beneficial. However, subcutaneous doses greater than 180 mcg in humans are limited by increasing drug toxicity, including significant liver transaminase elevations^21^. Finally, IFN-λ has been shown to disrupt the lung epithelial barrier in mice, leading to worsened disease course and increase susceptibility to bacterial superinfection^27,28^. This may negate any positive antiviral effects.

These data are in contrast to reports of benefits of Type I IFN in hospitalized COVID-19 patients. Subcutaneous IFN-β along with ribavirin and lopinavir/ritonavir was associated with shortened duration of symptoms and viral shedding in hospitalized patients in Hong Kong^14^, and a randomized clinical trial in England also suggest benefits of inhaled IFN-β for COVID-19.^29^ Although both type I and type III IFN activate the same dominant JAK-STAT signaling pathway^6^, inhibit SARS-CoV-2 *in vitro*^10,11^, and have receptors on respiratory epithelia^16^, *in vivo* activity and efficacy may differ^6^. A recent report found that inborn errors of Type I IFN immunity, including autosomal recessive IFNAR1 deficiency, were enriched in patients with life-threatening COVID-19 pneumonia^30^. Furthermore. patients with COVID-19 pneumonia were also more likely to have neutralizing auto-antibodies against type I, but not type III, IFNs^31^. These data suggest the possibility that type I IFN administration may be more beneficial than type III IFN in preventing adverse outcomes of SARS-CoV-2 infection.

Although there was some evidence that SARS-CoV-2 seropositivity at baseline modified the effect of treatment on shedding cessation, the effect modification was in the opposite direction than we had anticipated; Lambda appeared to prolong shedding relative to placebo among those who were seronegative at baseline, and to shorten the duration of shedding among those who were seropositive at baseline. Furthermore, Lambda also appeared to prolong shedding relative to placebo among those with high baseline viral RNA levels. These findings should be interpreted with caution for two reasons. First, these were exploratory analysis only and should be considered hypothesis generating. Second, these observations defy biological plausibility based on *in vitro* and animal model data.

The majority (62.5%) of participants in our study were Latinx, reflecting the high burden of COVID-19 among the Latinx community in our surrounding communities.^32^ Minority populations are disproportionately affected by COVID-19, with higher rates of infection and deaths observed due to a multitude of socioeconomic and demographic factors.^33,34^ Attention has recently been called to the relative absence of the most affected minorities in treatment trials^35,36^, and we prioritized recruitment efforts to the Latinx community in our study.

The study did have a few limitations. We recruited both symptomatic and asymptomatic patients. Asymptomatic patients contributed less to secondary outcomes since they presented with lower viral RNA levels and could not contribute to analyses of symptom alleviation. However, these patients represented <10% of the enrolled cohort. Additionally, despite a reported median duration of symptoms prior to randomization of only 5 days, 40% of participants were already seropositive at enrollment. Unpublished data from a Regeneron outpatient monoclonal antibody study with similar study design (REGN-COV2) found similar rates of baseline SARS-CoV-2 IgG seropositivity (45%).^37^ These data suggest that enrolling COVID-19 outpatients early in the course of disease, before they develop an antibody response, may be challenging. Nonetheless, we found no suggestion of benefit of Lambda in seronegative individuals. Finally, the median time to cessation in the placebo arm was shorter than assumed in our sample size calculations, potentially due to less severe disease in this population. However, our original sample size estimates based on the number of events and median time to event were conservative; a shorter time to cessation, keeping all other assumptions the same, increases the power to detect differences between groups.

In conclusion, a single dose of subcutaneous Peginterferon Lambda-1a, while safe, neither reduced time to cessation of viral shedding nor symptom duration in outpatients with uncomplicated COVID-19 in this large, Phase 2, single-center study. Further investigation into the therapeutic utility of type III interferons for COVID-19 in patients with severe illness or as a prophylactic treatment are underway.

## Methods

### Trial Design and Oversight

We conducted a Phase 2, single-blind, randomized placebo-controlled trial to evaluate the efficacy of Lambda in reducing the duration of viral shedding in outpatients. The trial was conducted within the Stanford Health Care System. Adults aged 18-65 years with an FDA emergency use authorized reverse transcription-polymerase chain reaction (RT-PCR) positive for SARS-CoV-2 within 72 hours from swab to the time of enrolment were eligible for participation in this study. Exclusion criteria included current or imminent hospitalization, respiratory rate >20 breaths per minute, room air oxygen saturation <94%, pregnancy or breastfeeding, history of decompensated liver disease, recent use of interferons, antibiotics, anticoagulants or other investigational and/or immunomodulatory agents for treatment of COVID-19, and prespecified lab abnormalities. Full eligibility criteria are provided in the study protocol. The protocol was amended on June 16^th^, 2020 after 54 participants were enrolled but before results were available to include adults up to 75 years of age and eliminate exclusion criteria for low white blood cell and lymphocyte count. The trial was registered at ClinicalTrials.gov (NCT04331899). The study was performed as an investigator initiated clinical trial with the FDA (IND 419217), and approved by the Institutional Review Board of Stanford University.

### Recruitment and Enrolment

Participants were recruited with flyers, online advertising, and phone calls to Stanford patients with positive SARS-CoV-2 RT-PCR. Recruitment materials and phone calls were provided in multiple languages, including English and Spanish. After confirming eligibility and providing informed consent in the patient’s primary language, participants underwent a standardized history and physical exam, and completed bloodwork. If inclusion criteria were met, participants were randomly assigned to Lambda or placebo using a 1:1 REDCAP-based computer-generated randomization scheme that stratified by age (≥ 50 and < 50 years old) and sex. A password-protected electronic spreadsheet containing the randomization allocation, along with the code used to generate the allocation and seed used in the random number generation, was stored on secure servers at Stanford.

### Study drug administration

Phase 2 studies established the optimal dose for virologic suppression and minimizing treatment-related adverse events (mainly aminotransferase and/or bilirubin elevations) for hepatitis C at 180 mcg given subcutaneously^21^. This dose is also currently being used in hepatitis D trials, and was provided by Eiger BioPharmaceuticals for use in this study. Those assigned to Lambda received a single 180 mcg subcutaneous injection of study drug (0.45 mL volume), and those assigned to placebo received a 0.45 mL subcutaneous injection of saline (prepared by the Stanford Investigational Pharmacy). The study medication/placebo syringe was dispensed by the Stanford Investigational Pharmacist and administered by a study nurse. Lambda and placebo syringes were identically labeled but differed in the appearance of the needle hub. Since the nurse administering the medication might see syringe differences, the study was not strictly “double-blind” even though all participants and investigators were blinded to treatment arm. Participants were monitored for adverse events for thirty minutes after injection.

### Participant Follow Up

Participants completed a daily symptom questionnaire using REDCap Cloud version 1.5. Participants also provided in-home measurements of temperature and oxygen saturation using study-provided devices. In-person follow-up visits were conducted at Day 1, 3, 5, 7, 10, 14, 21, and 28, with assessment of symptoms and vitals, and collection of oropharyngeal swabs (FLOQ Swabs; Copan Diagnostics). Peripheral blood was also collected at Day 5 and 14 to assess for safety events.

### Laboratory procedures

Laboratory measurements were performed by trained study personnel using point-of-care CLIA-waived devices or in the Stanford Health Care Clinical Laboratory. Oropharyngeal swabs were tested for SARS-CoV-2 in the Stanford Clinical Virology Laboratory using an emergency use authorized, laboratory-developed, RT-PCR.^38,39 40^ Centers for Disease Control and Prevention guidelines identify oropharyngeal swabs as acceptable upper respiratory specimens to test for the presence of SARS-CoV-2 RNA,^24^ and detection of SARS-CoV-2 RNA swabs using oropharyngeal swabs was analytically validated in the Stanford virology laboratory.

IgG antibody titers against the SARS-CoV-2 spike receptor binding domain (RBD) were assessed at enrolment.^25^ Briefly, heat inactivated serum samples at enrolment were diluted 5-fold starting at 1:50 and IgG antibody titers against RBD determined by ELISA. Absorbance was measured at 450nm (SPECTRAmax 250, Molecular Devices). Samples were considered seropositive against RBD if their absorbance value was greater than the mean plus four standard deviation (SD) of all negative controls (n=130).

### Data and Safety Monitoring

Adverse events were assessed and graded for severity according to standardized criteria.^20^ A Data and Safety Monitoring Board (DSMB) was established and conducted an interim analysis to review clinical trial progress, integrity, and safety data.

### Study Outcomes

The primary outcome was time to first of two consecutive negative oropharyngeal tests for SARS-CoV-2 by RT-PCR. Secondary outcomes included: 1) Time to alleviation of all symptoms, defined as time until the first day when no symptoms were reported; 2) SARS-CoV-2 oropharyngeal viral RNA levels over time; 3) SARS-CoV-2 oropharyngeal viral RNA area under the curve (AUC); and 4) Incidence of emergency department visits or hospitalizations within 28 days of initiation of treatment. Adverse events (AEs) and serious AEs (SAEs) were the primary safety endpoints. For secondary outcomes utilizing oropharyngeal viral RNA levels, we used the following conversion formula from cycle threshold values to copies/ml PBS:

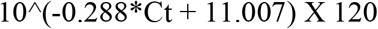

Exploratory outcomes included: 1) Time until sustained resolution of symptoms, defined as the first day when no symptoms were reported for the duration of the study; 2) Progression of disease, defined as admission to the emergency department, hospitalization, or worsening cough or shortness of breath defined as an increase in severity of two points or more on a five-point scale.

### Statistical Analysis

Analyses were performed according to assigned randomization arm (intent-to-treat). Absolute standardized differences are displayed to compare the distribution of baseline characteristics^41^. All models were covariate-adjusted for the randomization strata of age group and sex. Unless otherwise noted, all tests were two-sided and conducted at the 0.05 level of significance. Analyses were performed in R version 4.0.2.^42^

### Primary analysis

Time until shedding cessation was compared between arms using a Cox proportional hazards model covariate-adjusted for age and sex, with the final hypothesis test conducted at the alpha = 0·04999 level of significance to allow for an interim analysis. The hazard ratio for shedding cessation was estimated, along with its 95% confidence interval. Participants who dropped out prior to having two consecutive negative tests were censored at the time of their last positive test or on Day 1 if no positive tests were observed. The proportional hazards assumption was verified by examining the Schoenfeld residuals. Efron’s approximation was used to handle ties in the Cox proportional hazards model.

### Secondary analysis

Time until resolution of symptoms was compared using a Cox proportional hazards model. Hazard ratios and 95% confidence intervals were reported. Change in viral load during follow-up was compared using a linear mixed-effects model with random intercepts for participant. The AUC of viral load was compared using linear regression. Multiple imputation using chained equations was used to impute missing viral load data prior to area under the curve calculation. Five data sets were imputed, and imputed values calculated using non-missing viral load on each of the 7 sample collection days, treatment arm, age, sex, and whether or not a participant was hospitalized. Model estimates were pooled across the five imputed datasets by computing the total variance over the repeated analyses.

Estimates for change in viral load and viral load AUC for lambda compared to placebo and corresponding 95% confidence intervals for the linear models were reported. The incidence of hospitalizations and emergency department visits was estimated for each arm, with 95% confidence intervals. AEs were compared by arm using the Chi-squared test and Fisher’s exact test for SAEs.

A sensitivity analysis was performed for the primary endpoint using only symptomatic patients at baseline. Because two participants, after randomization, inadvertently were injected with the incorrect syringe, we also conducted an as-treated analysis according to treatment actually received.

A statistical interaction term between treatment arm and symptomatic status at baseline was added to the Cox proportional hazards model adjusted for age group and sex to test whether symptomatic status was an effect modifier of the relationship between treatment and time to shedding cessation. The main effect of each potential effect modifier was also included in the model. Additional effect modifiers specified a priori were 1) having a CT value < 30 (vs ≥ 30) on baseline oropharyngeal swab, 2) IgG seropositivity at baseline, and 3) number of risk factors or predictors for severe disease present at baseline (temperature ≥ 99.5, cough, or shortness of breath present at randomization [symptoms count as a single risk factor], age ≥ 60, male sex, Black race, Hispanic ethnicity, body mass index ≥ 30, and lab values of baseline lymphocyte counts < 1000 and baseline ALT ≥ 94). Effect modification was considered significant if the P value for interaction was <0.05.

Post hoc analyses were conducted to test for differences in both median duration of symptoms pre-randomization and baseline log_10_ viral load between seronegative and seropositive participants. The Kruskal-Wallis rank sum was used to test for differences in symptom duration while a two-sample t-test was used to test for differences in log_10_ viral load.

### Sample Size Determination

Assuming 1:1 randomization and the use of a two-sided log rank test at the alpha=0.04999 level of significance for the final analysis, we anticipated the occurrence of 79 shedding cessation events, which provided 80% power to detect a hazard ratio of 2.03. We additionally assumed median time to shedding cessation of 14 days in the control arm and 7 days in the treatment arm, a two-month accrual period, a two-week follow-up period after randomization of the last patient, and 10% drop out in the control arm. This enabled an interim analysis conducted at alpha=0·00001 to assess overwhelming efficacy after 50% of participants completed 24 hours of follow-up. We estimated that the total sample size required to achieve 79 events was 120 (60 participants per arm).

### Ethical approval

The study was registered as an investigator initiated clinical trial with the FDA (IND 419217), and approved by the Institutional Review Board of Stanford University. Written informed consent was provided by all study participants.

### Role of the funding source

The study was funded by anonymous donors to Stanford University, and Lambda provided by Eiger BioPharmaceuticals. The funders had no role in data collection and analysis or the decision to publish.

## Data Availability

Data will be made available at Stanford Digital Repository, lane.stanford.edu, at time of final publication.

## Acknowledgements

We thank all study participants who participated in this study, and the study team for their tireless work. We also thank our colleagues at Stanford University Occupational Health and at San Mateo Medical Center for participant referrals. Support from Stanford’s Innovation Medicine Accelerator and from Stanford ChEM-H is acknowledged. The Stanford REDCap platform (http://redcap.stanford.edu) is developed and operated by Stanford Medicine Research IT team. The REDCap platform services at Stanford are subsidized by a) Stanford School of Medicine Research Office, and b) the National Center for Research Resources and the National Center for Advancing Translational Sciences, National Institutes of Health, through grant UL1 TR001085.

## Authors’ Contributions

PJ, JA, JP, and US designed the study. PJ, JA, HB, KJ, SK, CV, OQ, KF, DW, JN, KE, CB, TW, BA, JP, and US collected data. CL, HH, JP, VB developed study data instruments. CH, IC provided study drug and guidance on study protocol. HH, NP, VB, and MD analyzed the data. All authors participated in data interpretation. KJ and PJ wrote the first draft and writing the manuscript and agreed on the decision to publish. There were no confidentiality agreements between the sponsors and authors.

## Data Availability Statement

The datasets generated and analyzed during the current study will be made available in the Stanford Digital Repository (lane.standord.edu) at the time of publication.

## Declaration of Competing Interests

Colin Hislop and Ingrid Choong are scientists at Eiger BioPharmaceuticals, Inc., which provided the Interferon Lambda used for this study. Jeffrey Glenn serves on the board of Eiger BioPharmaceuticals, Inc. Colin Hislop and Ingrid Choong own stock and options of Eiger BioPharmaceuticals, Inc. Jeffrey Glenn has an equity interest in Eiger BioPharmaceuticals, Inc. Jeffrey Glenn and Ingrid Choong are inventors on a pending patent application relating to the use of interferon lambda for coronavirus. Eiger BioPharmaceuticals played no role in study design, conduct of the study, or analysis of the data.

## Notes

### Clinical Trial

ClinicalTrials.gov NCT04331899

### Author Declarations

Stanford University Institutional Review Board

