## Supplemental for "Peginterferon Lambda-1a for treatment of outpatients with uncomplicated COVID-19: a randomized placebo-controlled trial"

Supplemental Appendix

### Supplementary Table 1. Exploratory Effect Modifier Results for Primary Outcome (Viral Cessation)

| **Effect modifier** | **Interaction p-value** |  | **Median time (95% CI) to shedding cessation** | | **Hazard ratio (95% CI) for lambda vs placebo** |
| --- | --- | --- | --- | --- | --- |
|  |  | **Subgroup** | **Lambda** | **Placebo** |  |
| Ct value < 30 | 0.15 | Ct < 30 | 15 (14, undefined) | 11 (10, 14) | 0.51 (0.26-1.04) |
|  |  | Ct 30+ | 3 (3, 7) | 5 (2, 7) | 0.95 (0.60, 1.52) |
| Seropositivity | 0.03 | Seronegative | 13 (7, 21) | 10 (7, 13) | 0.66 (0.39, 1.10) |
|  |  | Seropositive | 1 (0, 7) | 5 (2, 10) | 1.58 (0.88, 2.86) |
| Risk score | 0.77 | NA | NA | NA | 1.05 (0.76, 1.45) |
| Age 50+ | 0.21 | Age < 50 | 7 (5, 14) | 7 (5, 10) | 1.20 (0.59, 2.47) |
|  |  | Age 50+ | 3 (3, 14) | 7 (5, 21) | 0.70 (0.44, 1.10) |
| Male | 0.74 | Female | 7 (3, 14) | 5 (5, 12) | 0.86 (0.52, 1.42) |
|  |  | Male | 7 (5, 14) | 10 (7, 11) | 0.75 (0.42, 1.36) |

CT: Cycle Threshold by real time quantit:

Risk score is defined as the number of relevant severe disease risk factors present at baseline (presence of either temperature

of 99.5F+, cough, or shortness of breath; age 60+; male sex; Black race; Hispanic ethnicity; BMI 30+; ALC<1000; ALT 94+).

Hazard ratio >1 favors faster shedding cessation in Lambda vs. placebo arms; hazard ratio <1 favors delayed shedding cessation in Lambda vs. placebo arms. All models adjusted for age group and sex.

### Supplemental Figure 1. Symptom duration at presentation

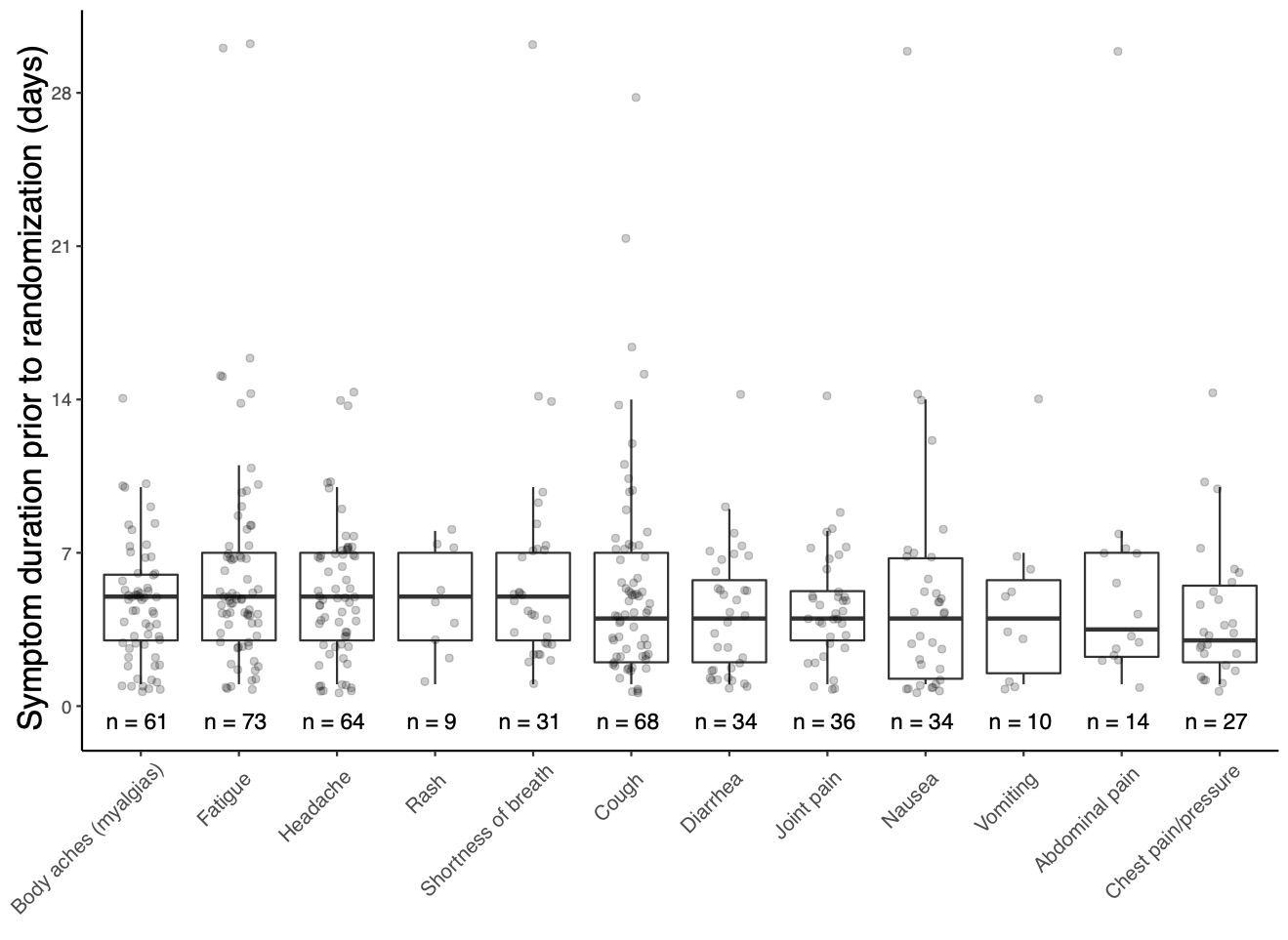

**Supplemental Figure 1.** Symptom duration at presentation

### Supplemental Figure 2. Time until SARS-CoV-2 viral shedding cessation from oropharyngeal swabs stratified by baseline SARS-CoV-2 IgG seropositivity

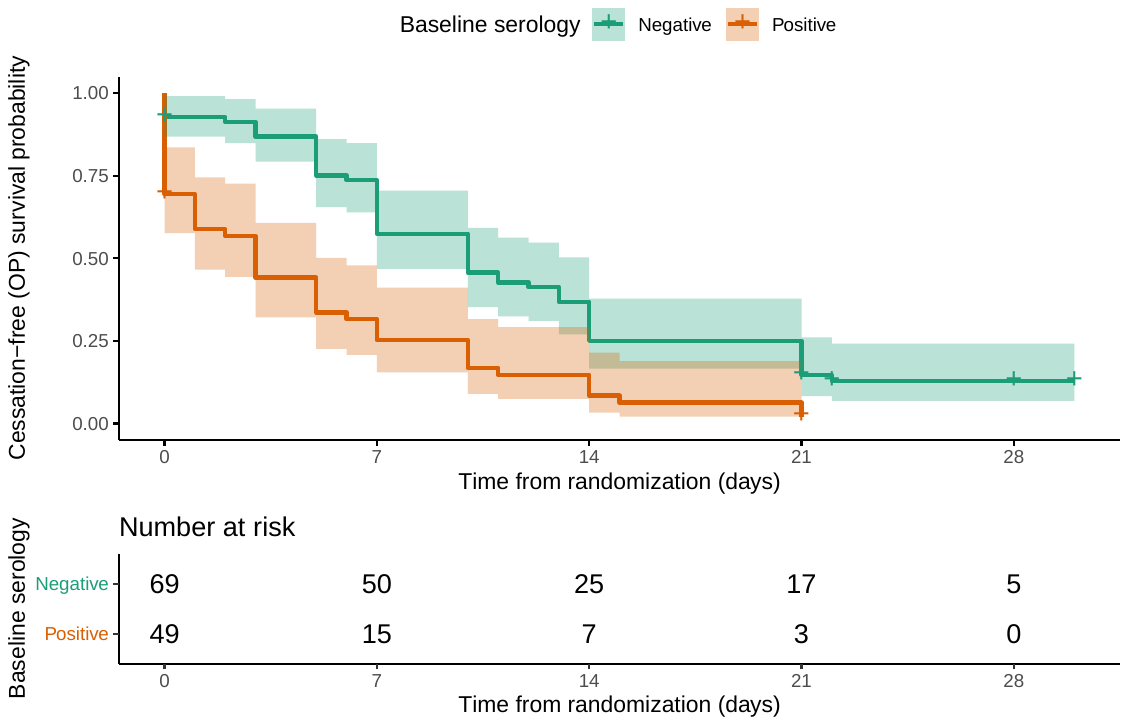

**Supplemental Figure 2.** Kaplan-Meier Analyses of the time until cessation of SARS-CoV-2 viral shedding from oropharyngeal swabs stratified by baseline SARS-CoV-2 seropositivity, Seronegative (green) vs. Seropositive (red).

### Supplemental Figure 3. Time until SARS-CoV-2 viral shedding cessation from oropharyngeal swabs stratified by baseline oropharyngeal viral load

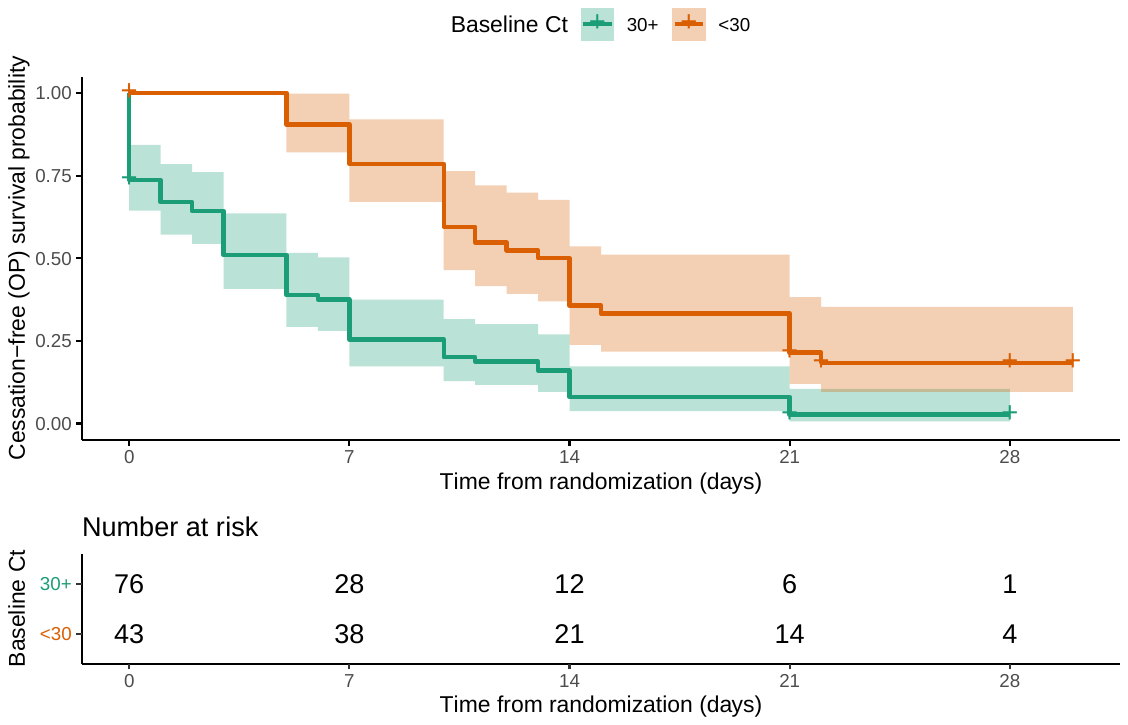

**Supplemental Figure 3.** Kaplan-Meier Analyses of the time until cessation of SARS-CoV-2 viral shedding from oropharyngeal swabs stratified by baseline oropharyngeal SARS-CoV-2 cycle threshold, >30(green) vs. <30 (red).

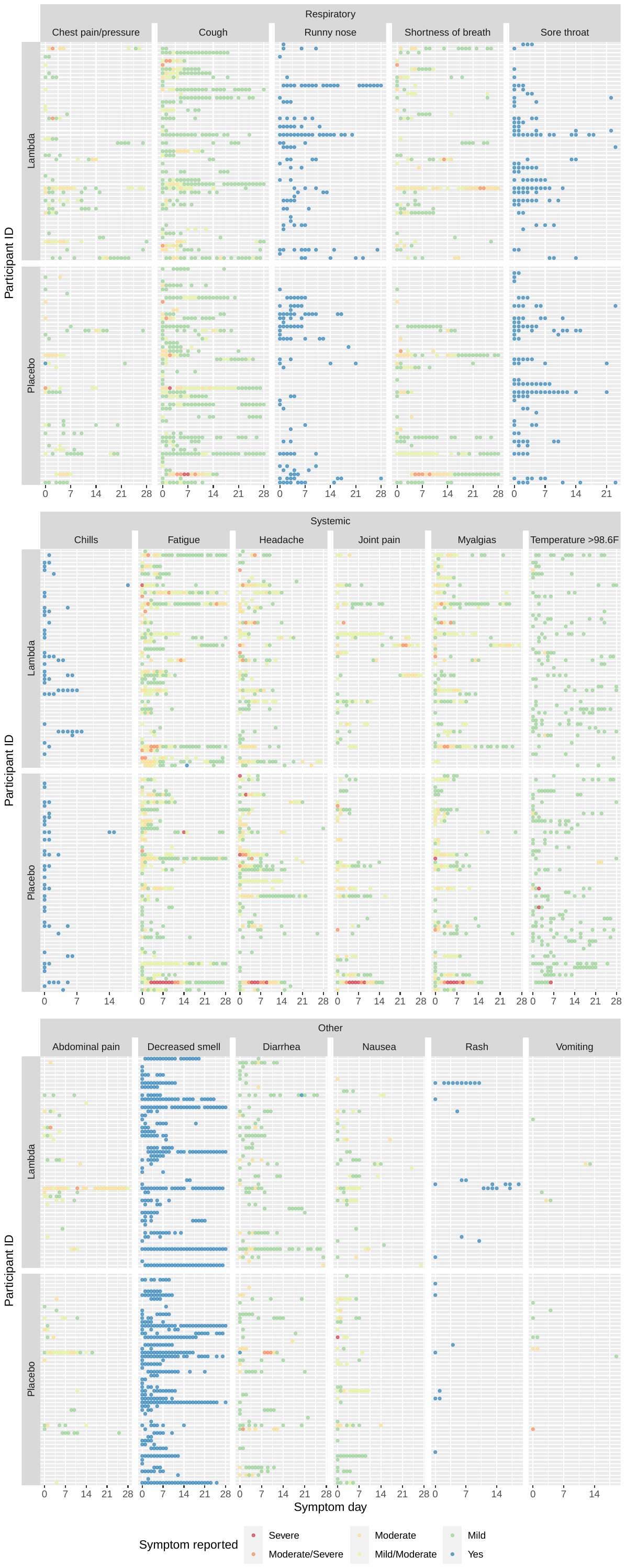

### Supplemental Figure 4. Individual participant symptoms over course of study, stratified by symptom complex and treatment arm.

**Supplemental Figure 3.** Individual participant symptoms, stratified by symptom complex (respiratory, systemic, or other) and treatment arm. Colors represent severity of symptom reported, or symptom presence (blue circles) for those symptoms where severity was not assessed.

### Supplemental Figure 5. Oropharyngeal viral load over time

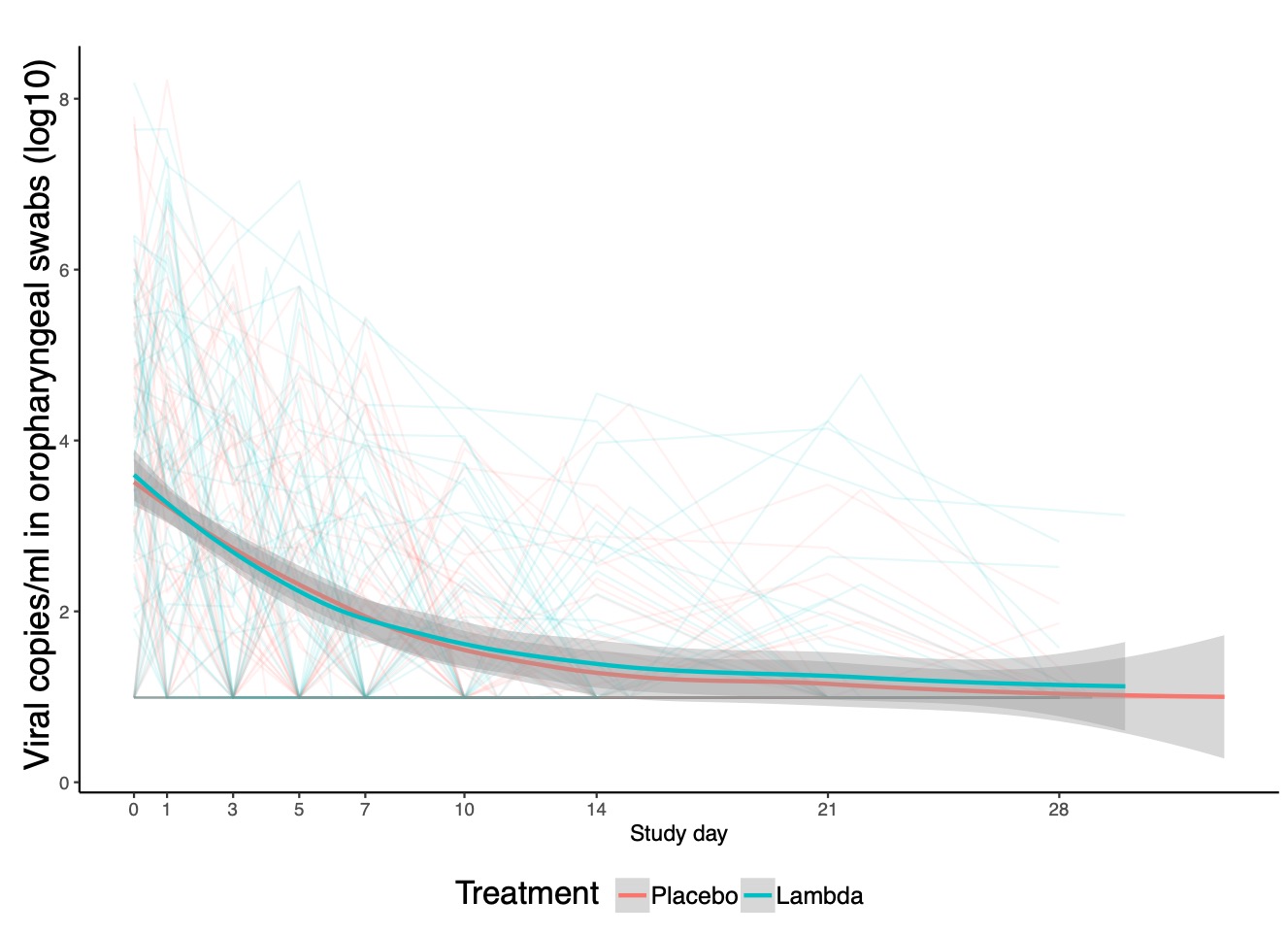

**Supplemental Figure 2.** Viral copies/ml in oropharyngeal swabs over course of study, by participant and treatment arm. Shown are LOESS regression lines with 95% CI, stratified by treatment arm.

### Appendix 1: Daily symptom questionnaire

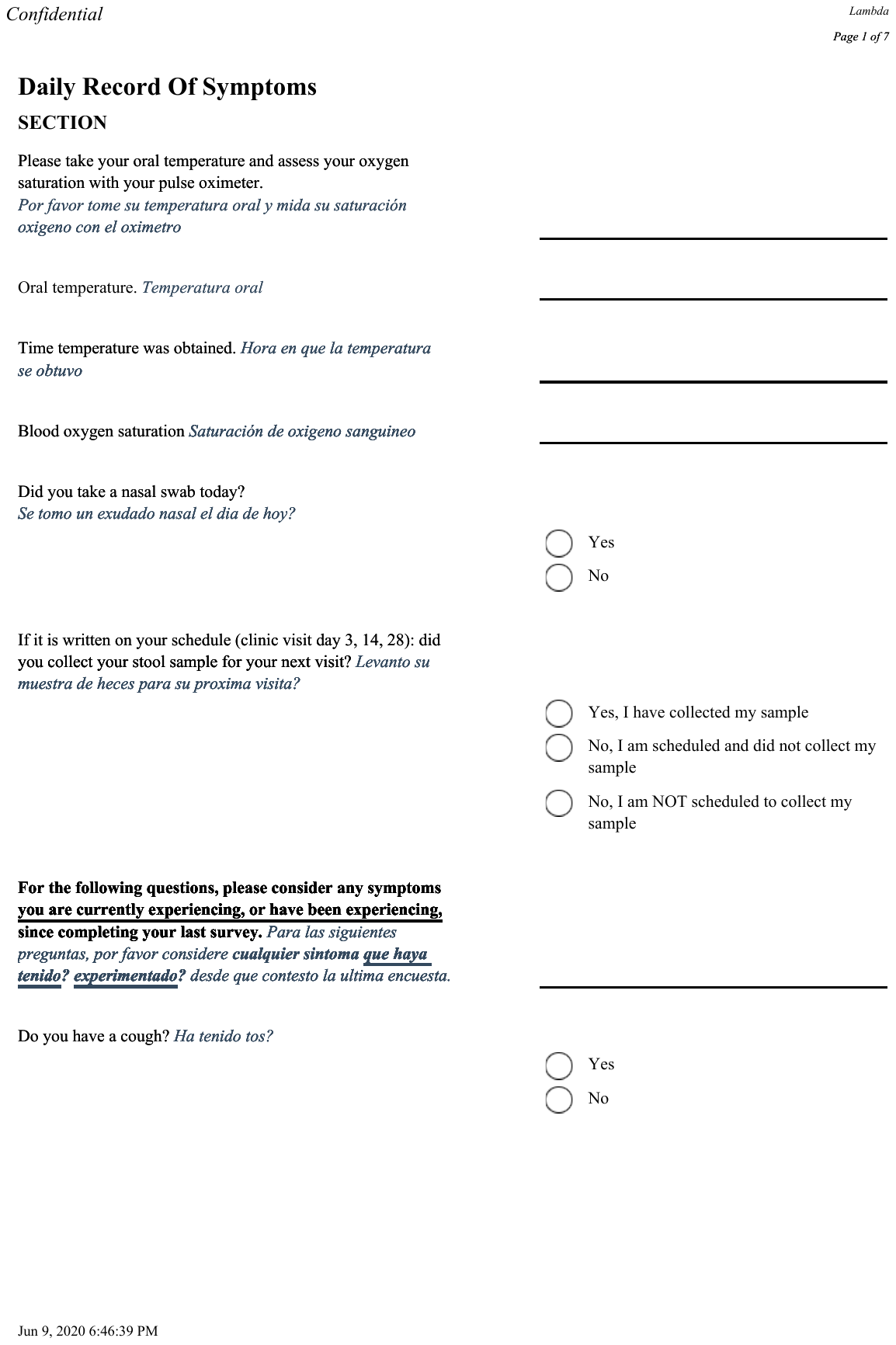

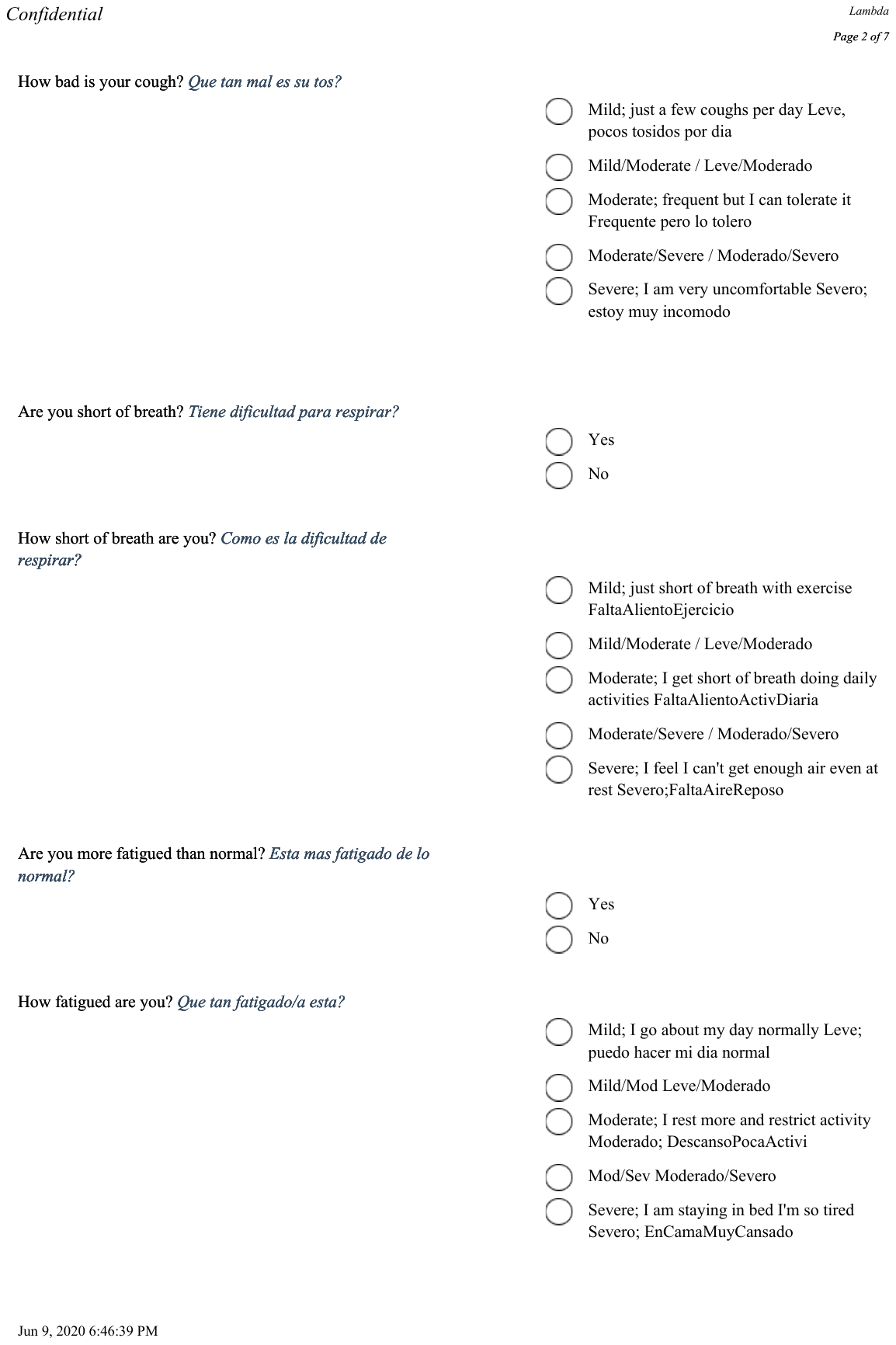

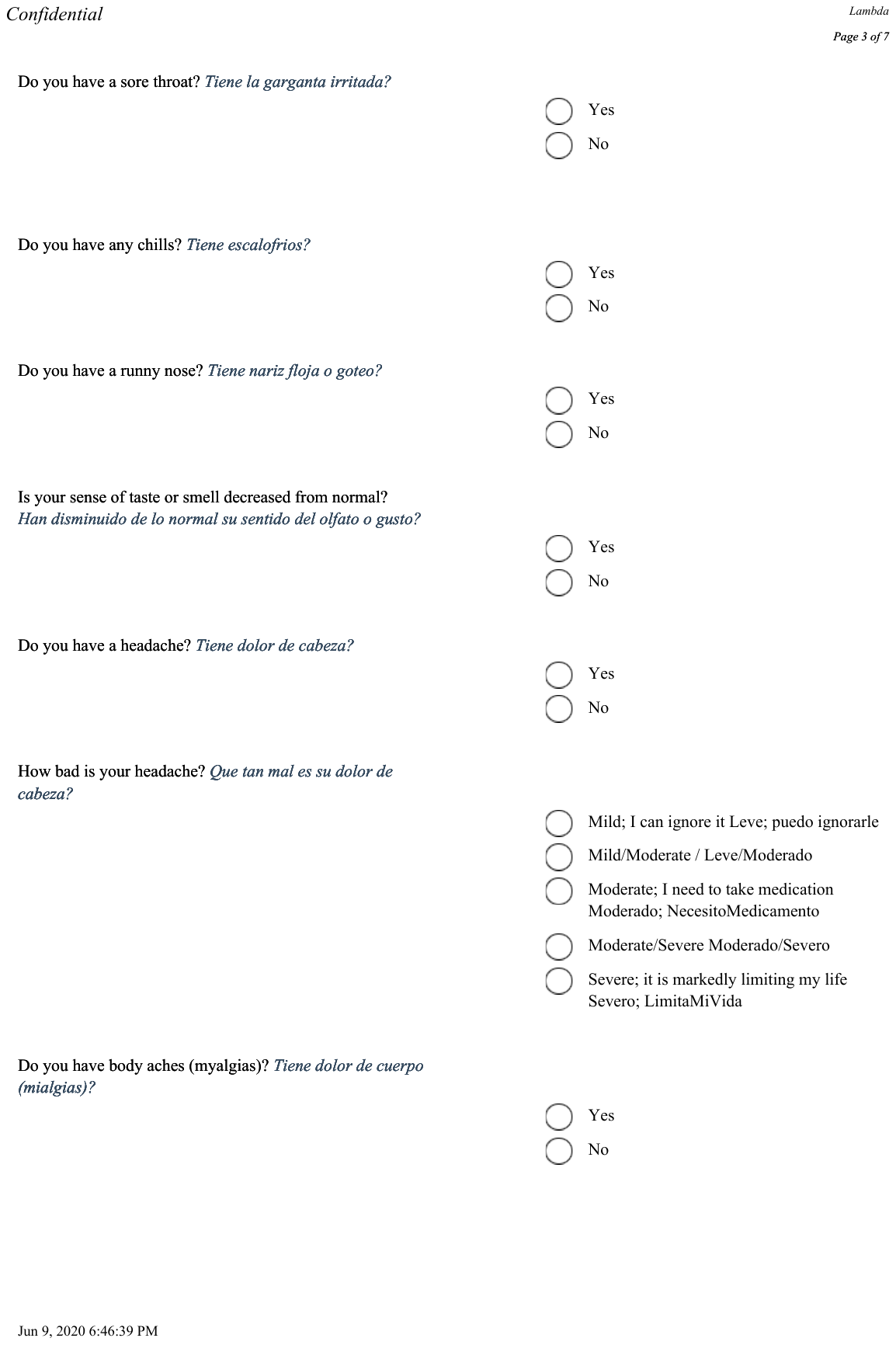

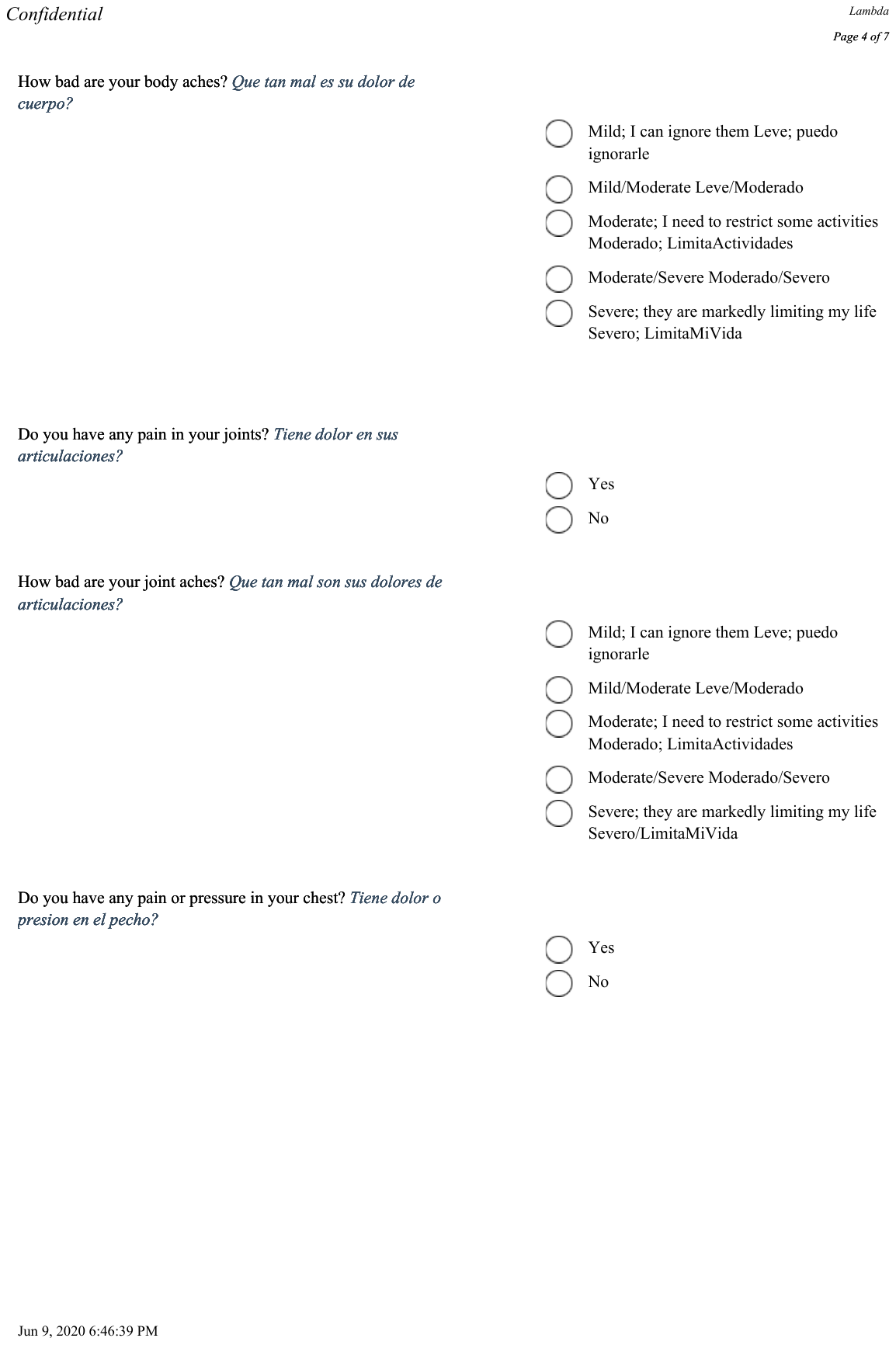

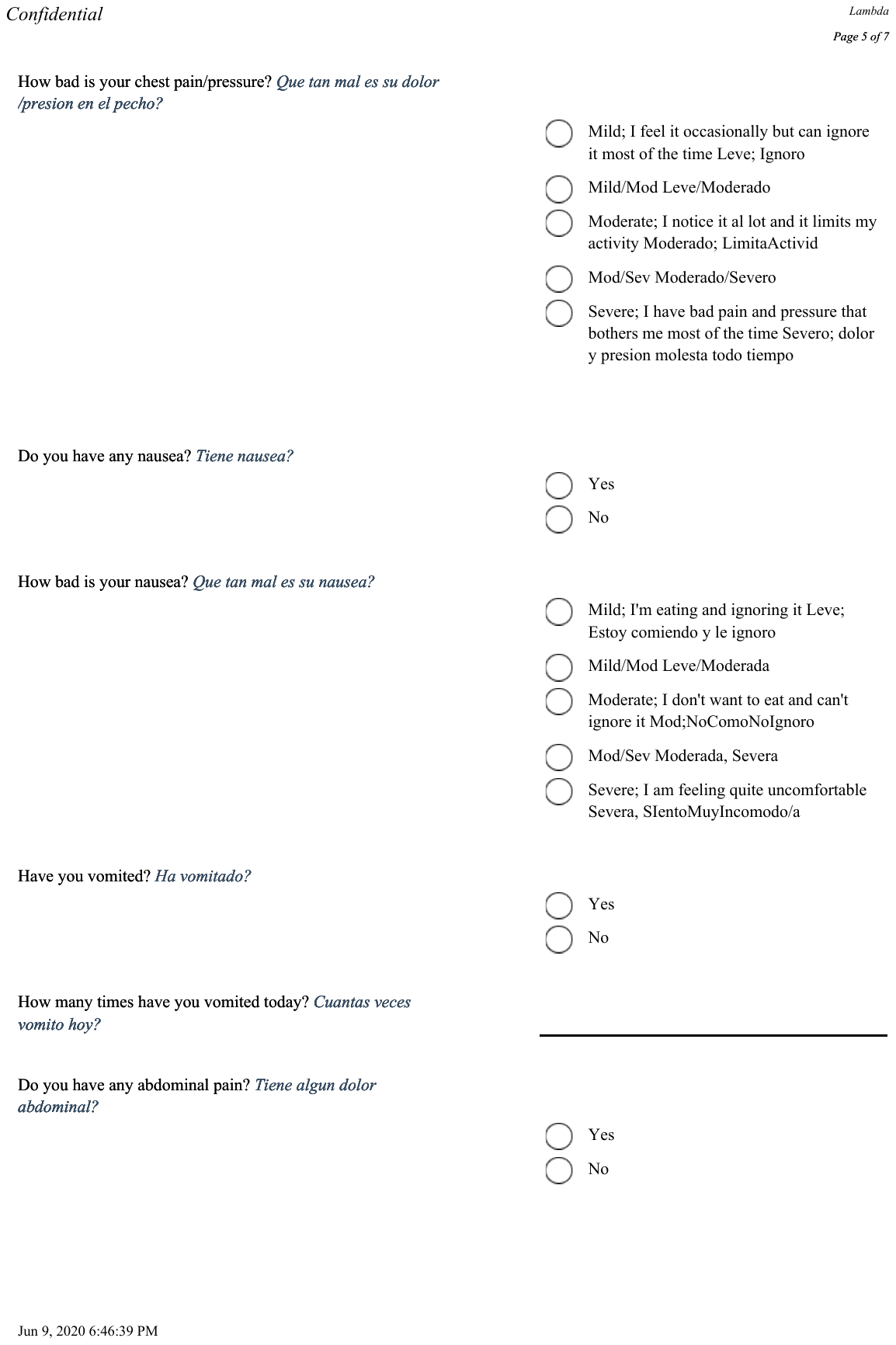

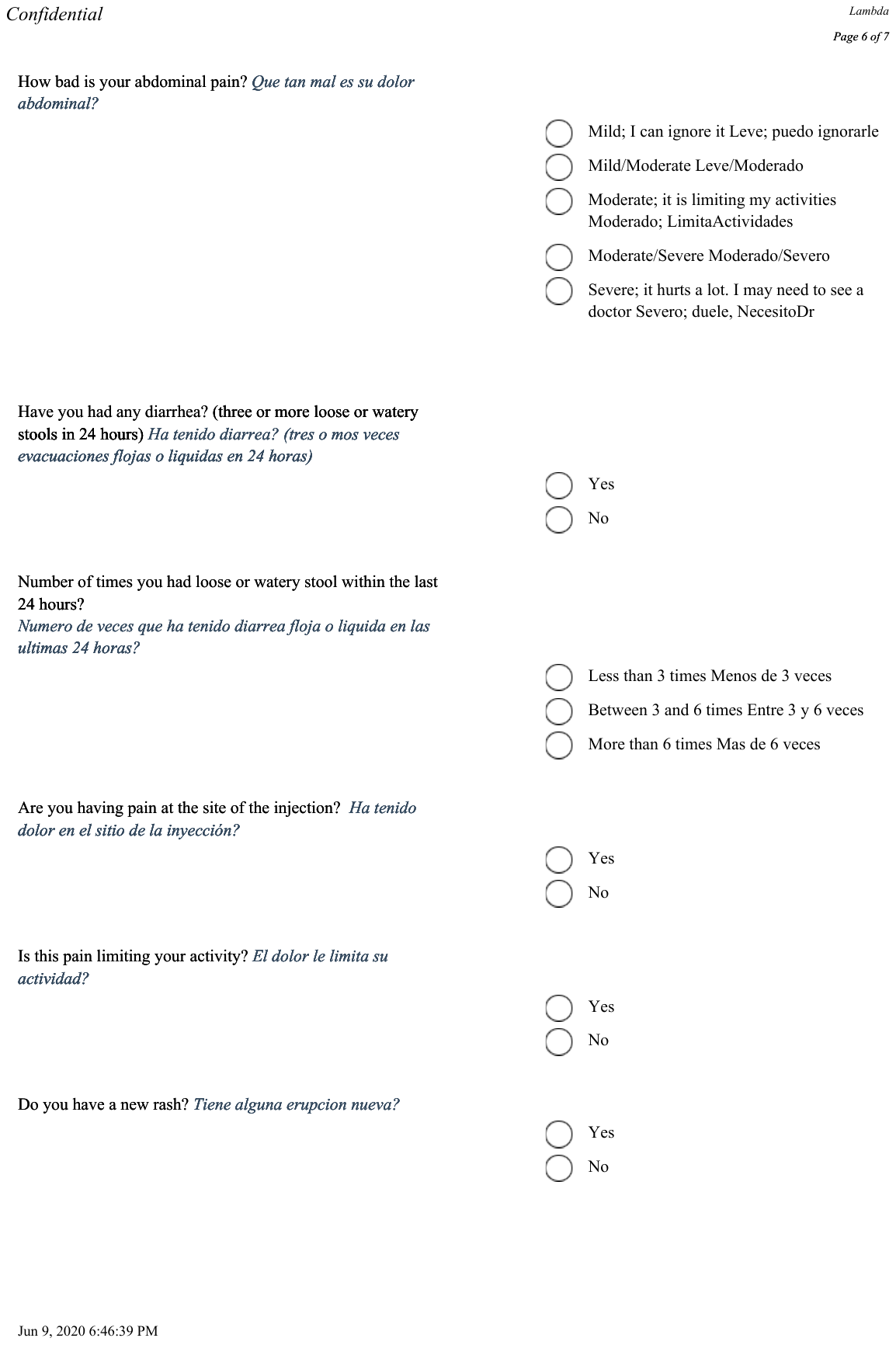

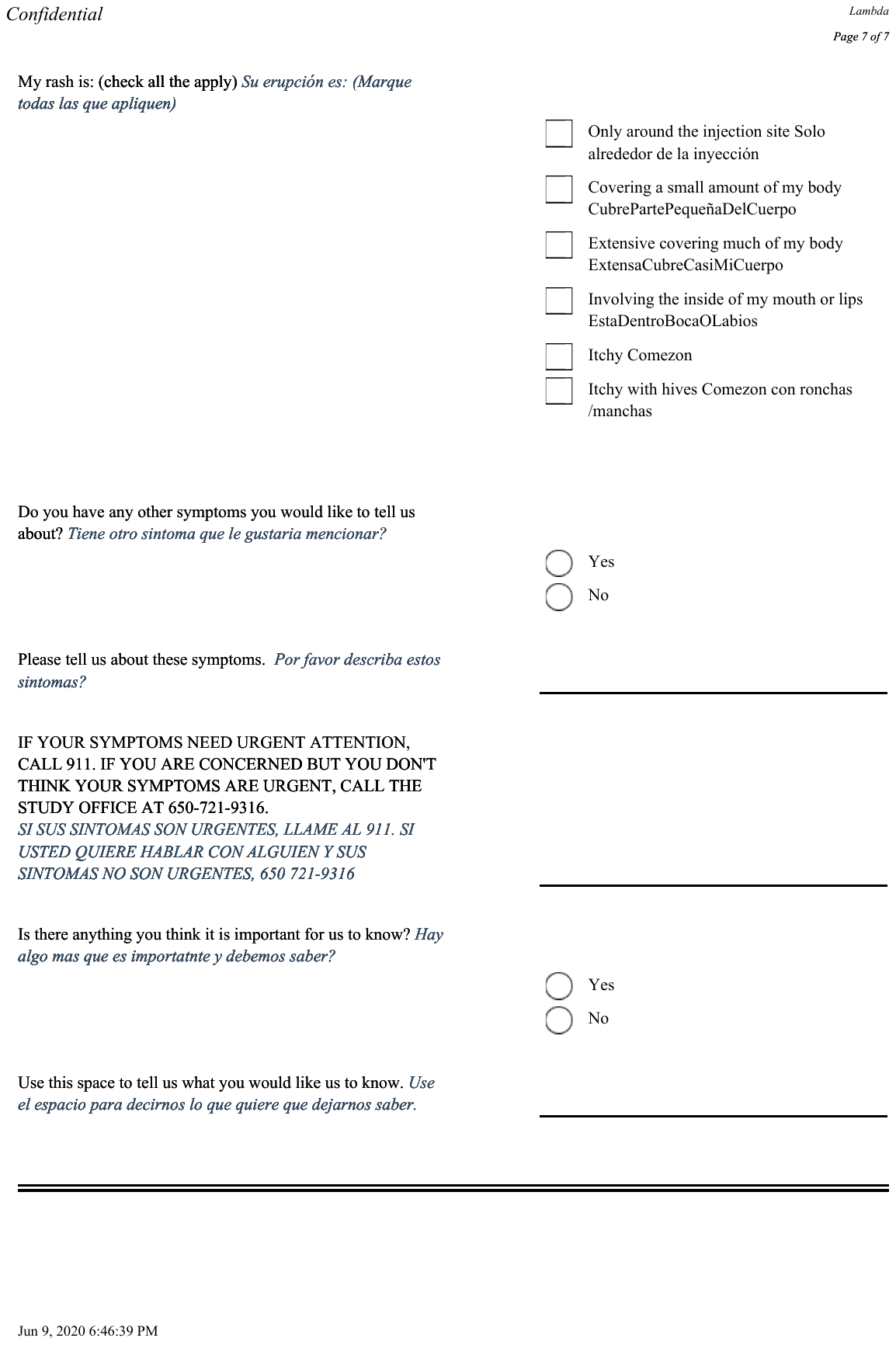
